# Pandemic trends in health care use: From the hospital bed to the general practitioner with COVID-19

**DOI:** 10.1101/2021.07.09.21260249

**Authors:** Fredrik Methi, Kjersti Helene Hernæs, Katrine Damgaard Skyrud, Karin Magnusson

## Abstract

**Aim:** To explore whether the acute 30-day burden of COVID-19 on health care use has changed from the beginning to the end of the pandemic.

**Methods:** In all Norwegians (N=122 699) who tested positive for SARS-CoV-2 in three pandemic waves (March 1^st^-July 31^st^ 2020 (1^st^ wave), August 1^st^-December 31^st^ 2020 (2^nd^ wave), and January 1^st^-May 31^st^ 2021 (3^rd^ wave)), we studied the age- and sex-specific share of patients (by age groups 1-19, 20-67, and 68 or more) who had: 1) Relied on self-care, 2) used primary care, and 3) used specialist care.

**Results:** We find that a remarkably high and stable share (70-80%) of patients with COVID-19 exclusively had contact with primary care in the acute phase, both in the 1^st^, 2^nd^ and 3^rd^ wave. The mean number of primary care visits ranged between 2 and 4. We also show that the use of specialist care in the acute 30-day phase of COVID-19 has decreased, from 14% being hospitalized at least once during spring 2020, to 4% during spring 2021. The mean number of hospital bed-days decreased significantly for men from the 1^st^ to the 2^nd^ wave (from 13 days, 95% CI=11.5-14.5 to 10 days (9-11) for men aged ≥68 years, and from 11 days (10-12) to 9 days (8-10) for men aged 20-67 years), but not for women.

**Conclusion:** COVID-19 places a continued high demand on the primary care services, and a decreasing demand on the specialist care services.

## Introduction

The World Health Organization declared the outbreak of COVID-19 a global pandemic on 11 March 2020 [1]. Since then, most countries have implemented strict lockdown measures in order to control the virus and limit the burden to the health care system. The goal of lockdown has been to ensure a sufficient capacity in inpatient specialist care, i.e. enough hospital beds and ventilators had to be available when large proportions of the population were infected. The clinical course and mortality of COVID-19 among hospitalized individuals have therefore been well described, especially for the first wave of the pandemic (spring 2020) [2-5].

Less is known about COVID-19’s total use of health care services, including both specialist and primary care, both during the 1^st^ wave, but especially during the 2^nd^ and 3^rd^ wave striking fall 2020 and spring 2021 in Norway. To date, studies of primary care use during the pandemic have focused on structural changes in its delivery, such as telemedicine visits vs. office-based visits [6], whereas studies of the use of primary care services are lacking. The use of health care services may be hypothesized to have changed throughout the pandemic, starting with limited test availability and many persons in risk groups being hospitalized and eventually dying, to mass testing, mass vaccination of persons at risk and an increasing herd immunity. In the latest wave, the health care services may be hypothesized to be better trained in how to manage severely ill patients, leading to declining specialist care treatment and fewer fatal outcomes today than in the beginning of the pandemic.

Describing how the acute impact of COVID-19 on primary and specialist care services has behaved from the 1^st^ to the 3^rd^ wave of infection, is important when trying to predict how the burden on health care services will be in a 4^th^ or 5^th^ wave. It is also important for the understanding of how a similar future pandemic in its different phases would affect the health services, given everything we have learnt from the implementation of lockdown measures. More specifically, for the timely and correct upscaling or downscaling of the health services, we need to understand the major COVID-19 patient flows through the health care systems, as well as the peak and total demand of health care services in the days following a positive test of an individual.

In this paper we aim to explore the age- and sex-specific acute burden of COVID-19 on the health care services in three waves of the pandemic in Norway. Starting with the date for the first positive test for SARS-CoV-2, we estimated the wave-specific major patient pathways, as well as the peak and total use of health care services, for children and adolescents (age 1-19 years), the working age population (20-67) and the elderly (68 and above) within a 30-day time frame.

## Methods

The BeredtC19-register is an emergency preparedness register aiming to provide rapid knowledge about the pandemic, including impacts of measures to limit the spread of the virus on health and utilization of health care services [7]. BeredtC19 compiles daily updated individual-level data from several registers. It includes the Norwegian Surveillance System for Communicable Diseases (MSIS) (all testing for COVID-19), the Norwegian Patient Register (NPR) (all electronic patient records from all hospitals in Norway), and the Norway Control and Payment of Health Reimbursement (KUHR) Database (all consultations with all general practitioners and emergency primary health care), as well as the National Population Register (age, sex, country of birth, date of death). Thus, the register includes all polymerase chain reaction (PCR) tests for SARS-CoV-2 in Norway with date of testing and test result, reported from all laboratories in Norway and all electronic patient records from primary care as well as outpatient and inpatient specialist care. The establishment of an emergency preparedness register forms part of the legally mandated responsibilities of The Norwegian Institute of Public Health (NIPH) during epidemics. The Ethics Committee of South-East Norway confirmed (June 4th 2020, #153204) that external ethical board review was not required.

### Population

Our population included every Norwegian resident on January 1st 2020 who tested positive for the SARS-CoV-2 by a PCR-test from March 1^st^ 2020 to May 31^st^ 2021. The date with the first record of a confirmed test was coded as being the start of the individual’s health care pathway. Patients with negative PCR-tests, as well as patients with suspected COVID-19 and without positive PCR-tests were excluded. We divided our population into mutually exclusive age and sex groups, i.e. girls and boys, men and women by the following age categories: 1-19 (children and adolescents), 20-67 (working age population) and 68 years or older (elderly).

### Outcomes

We had several outcomes, which combined, and sorted chronologically on dates of occurrence provided a comprehensive picture of COVID-19-related health care pathways. The dates of all outcomes were sorted relative to the date of first positive PCR-test, with the test date being coded as day 0 and the outcomes occurring on day 0 to day 30. To take into account that many PCR-tests were prescribed by the health care services, we performed analyses that included vs. not included visits related to the testing. Our outcomes were defined as follows:

1. Primary care use with International Classification of Primary Care (ICPC-2) code R992 (COVID-19) (GPs or emergency wards).
2. Hospital-based inpatient specialist care with International Classification of Disease (ICD-10) code U071 (confirmed COVID-19).
3. Death independent of cause but occurring within 30 days after the positive test.

## Statistical analyses

We first assessed descriptive statistics of our study population by different waves or periods of the pandemic, each having different characteristics: the 1^st^ wave of transmission (March 1^st^ 2020 – July 30^th^ 2020) which was characterized by low availability of testing (only available for health personnel, elderly and persons at risk), the 2^nd^ wave of transmission (August 1^st^ 2020-December 31^st^ 2020), which was characterized by wide testing criteria and free testing, and the 3^rd^ wave of transmission (January 1^st^ 2021 – June 1^st^ 2021), which was characterized by continued wide testing criteria and free testing as well as the start of mass vaccination and an increasing number of the more transmissible variants (especially alpha, but also some cases of beta, gamma and delta) [8-10].

Second, based on the total health care use observed during the -2 to +30 days following positive test, we divided the study population into three different major patient pathways. Each of the patient pathways represented a different acute burden of disease on the health care systems: 1) patients who had no contact with any health services, 2) patients who had contact with primary care (GP and/or emergency ward) only, and 3) patients who had contact with specialist care, with or without additional need for primary care. To get an overarching picture of the major patient flows, we visualized the timing of care for these different pathways in alluvial diagrams (see S-Figure 1). We also estimated the whole-sample- and pathway-specific mortality as proportions with 95% confidence intervals using the Wilson-method (see S-Table 1).

Third, we studied the peak and total use of primary and specialist care during the -2 to +30 days following positive test, for each of the 1^st^, 2^nd^ and 3^rd^ waves, independent of whether the care use could be classified in one of the above-described care pathways. To explore the wave-wise *peak use*, we estimated day-by-day proportions that visited primary and specialist care at least once during day -2 to +30. For persons with at least one visit in primary care, we included the first visit, and for persons with at least one visit in specialist care, we coded all the days spent in hospital as hospital bed-days. To explore the *total use* in each wave, we estimated the cumulative proportions visiting primary or specialist care at least once during day -2 to +30. We repeated these analyses with exclusion of day –2 to 0 to account for the impact of testing on the peak and total use of health care. Finally, among persons having at least one visit in primary or specialist care, respectively, and by age and sex, we also estimated the mean number of visits in primary care and mean number of days spent in hospital (bed-days) in each wave (with 95% confidence intervals being calculated as 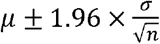 where µ is the mean, σ is the standard deviation and n is the population size). All analyses were run using STATA SE v.16.

## Results

We identified 122 699 persons with at least one positive PCR-test for SARS-CoV-2 in the total tested population of 3 096 200 persons between March 1^st^ 2020 and May 31^st^ 2021. The total number of tests throughout the wave was 6 918 216. The percentages positive tests among all tests in the 1^st^, 2^nd^ and 3^rd^ waves were 1.9%, 1.6% and 2.2%, respectively. Table 1 shows that the age of persons testing positive decreased from the 1^st^ to the 3^rd^ wave. It also shows that the percentage of women among those testing positive decreased from the 1^st^ to the 3^rd^ wave (Table 1). The proportions dying within 30 days after positive test decreased from the 1^st^ to the 2^nd^ wave (i.e. from 18% to 8% for men above 68 years and from 16% to 9% for women above 68 years), but not or to a lesser extent from the 2^nd^ to 3^rd^ wave (S-Table 1). The 30-day mortality was low across all pandemic waves for persons aged under 67 years (S-Table 1).

**Table 1.** Descriptive characteristics of persons testing positive for SARS-CoV-2 in each of three pandemic waves in Norway, 2020-2021.

|  | 1st wave<br>March 1st -<br>July 31st 2020<br>N=9158 | 2nd wave<br>August 1st -<br>December 31st<br>2020<br>N=39 595 | 3rd wave<br>January 1st -<br>May 31st 2021<br>N=73 946 |
| --- | --- | --- | --- |
| Age, mean (SD) | 45.6 (19.7) | 36.0 (19.1) | 31.5 (18.4) |
| Women, N (%) | 4590 (50.1) | 18621 (47.0) | 34460 (46.6) |
| Children and adolescents (1-19 years) |  |  |  |
| Girls, N (%) | 330 (3.6) | 3988 (10.1) | 11048 (14.9) |
| Boys, N (%) | 333 (3.6) | 4197 (10.6) | 11716 (15.8) |
| Adults in working age (20-67 years) |  |  |  |
| Women, N (%) | 3604 (39.4) | 13290 (33.6) | 22202 (30.0) |
| Men, N (%) | 3616 (39.5) | 15574 (39.3) | 26544 (35.9) |
| Elderly ($\geq 68$ years) | | | |
| Women, N (%) | 656 (7.2) | 1343 (3.4) | 1210 (1.6) |
| Men, N (%) | 619 (6.8) | 1203 (3.0) | 1226 (1.7) |

### Patient pathways from positive test to 30 days after

Boys and girls aged 1-19 years largely followed the same patient pathways in all three waves, whereas men aged ≥20 years were more frequently treated in specialist care compared to women aged ≥20 years who were more frequently treated in primary care (Figure 1).

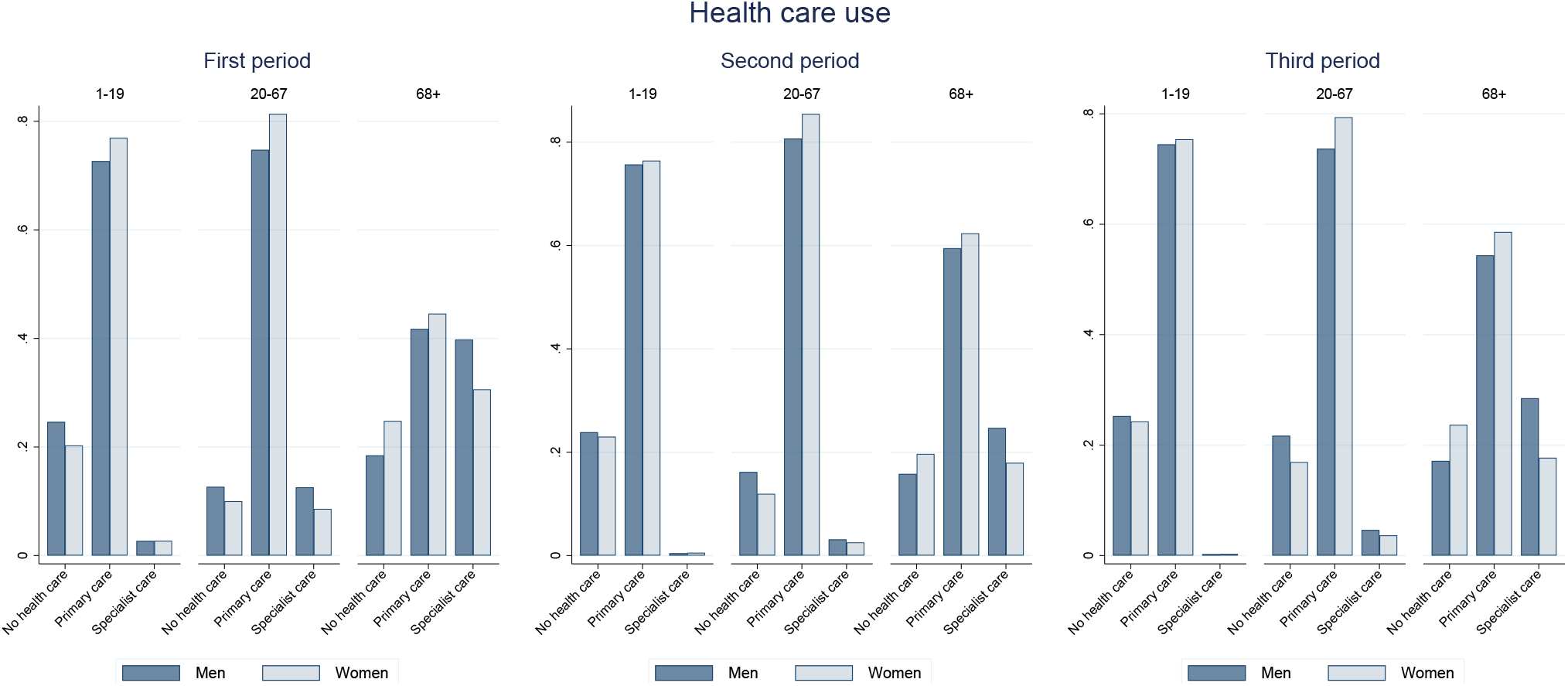

The share that did not use health services was relatively stable across the three waves for boys and girls aged 1-19 years as well as for the elderly (68 or more) (Figure 1). For men and women in their working age (20-67) the share relying on self-care increased from the 1^st^ to the 3^rd^ wave, especially for women (Figure 1).

The share using only primary care was stable across the waves, around 75-85% for persons aged 1-67 (Figure 1). In contrast, for the elderly, the share using only primary care increased from ∼40% in the 1^st^ wave, to ∼60% in the 2^nd^ and 3^rd^ wave (Figure 1). Along these shifts, we observed a decrease in the share of persons needing specialist care, especially from the 1^st^ to the 2^nd^ wave, but not from the 2^nd^ to 3^rd^ wave, for all groups of age and sex (Figure 1). Further, the 30-day all-cause mortality decreased for persons in all healthcare pathways from the 1^st^ to the 3^rd^ wave (S-Table 1). However, women aged ≥68 years who were treated in specialist care had a less varying mortality across the three waves (S-Table 1).

S-Figure 1 shows that the share in need of specialist care prior to primary care was larger than the share in need of primary care prior to specialist care, for all pandemic waves. Furthermore, the shares that were still in need of primary or specialist care on day 30 after a positive test decreased from the 1^st^ to the 2^nd^ wave and to a lesser extent from the 2^nd^ to the 3^rd^ wave, for all groups of age and sex. During the 1^st^ wave, 60% of persons aged ≥68 years did not use health care services on day 21-30 after a positive test, compared to 80% during the 3^rd^ wave (S-Figure 1).

### Peak and total use of primary care

Figure 2 shows a small decline in both the peak and cumulative use of primary care, from the 1^st^ to the 2^nd^ and to the 3^rd^ wave. During the 1^st^ wave, half of all patients visited primary care on day 0, decreasing to 40% in the 2^nd^ wave and 25% in the 3^rd^ wave (Figure 2). Furthermore, during the 1^st^ wave, patients continued to visit primary care for a longer time compared to the other waves, with still 2-5% visiting primary care during days 20 and 30 (Figure 2). S-Figure 2 shows that middle-aged and elderly persons continued to visit primary care for a longer time than persons aged 1-19 years. For adults aged 20 or more, the mean number of visits in primary care decreased from the 1^st^ to the 3^rd^ wave in the pandemic, whereas it increased for children and adolescents aged 1-19 years (Figure 3). When we excluded primary care use in relation to testing, results showed a slight increase in the cumulative share from the 1^st^ wave (72%) to the 2^nd^ wave (76%), and a slight decrease again in the 3^rd^ wave (74%) (S-Figure 3).

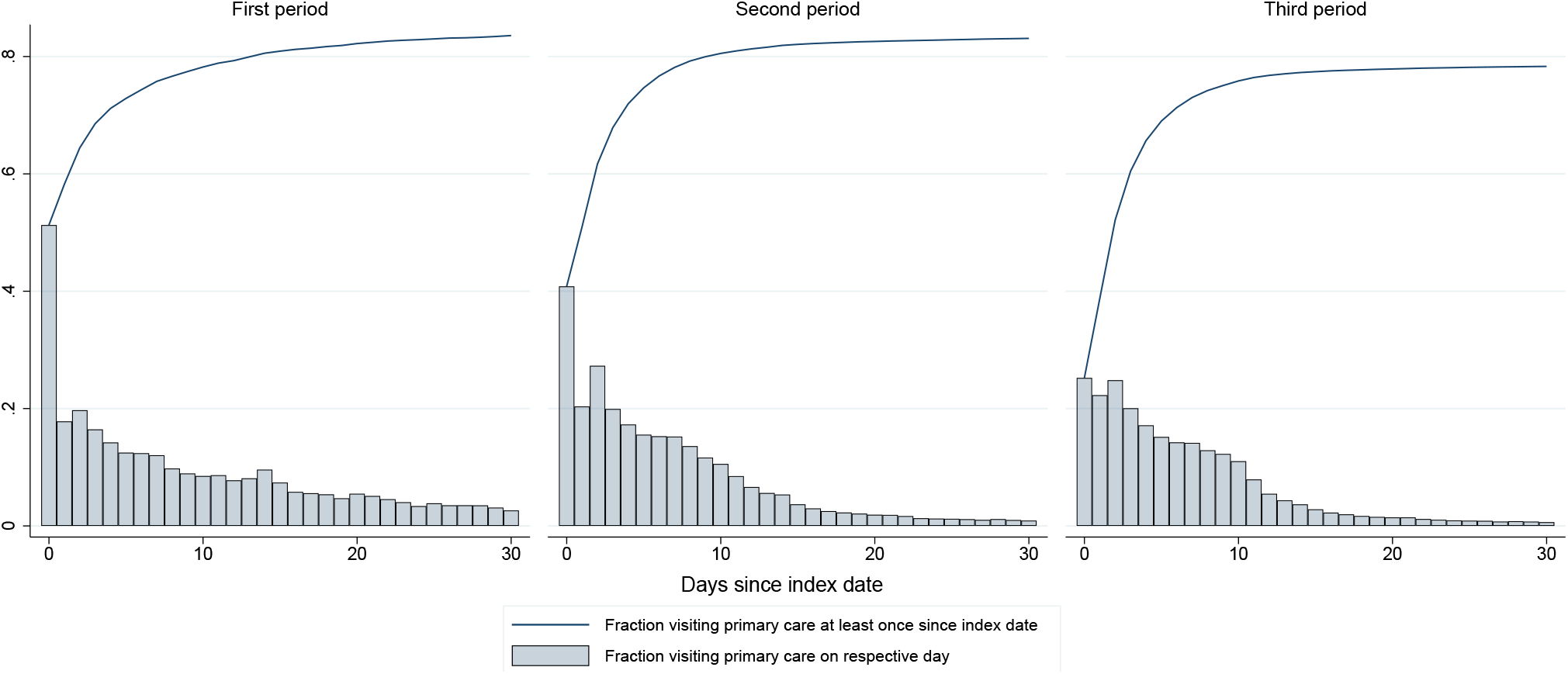

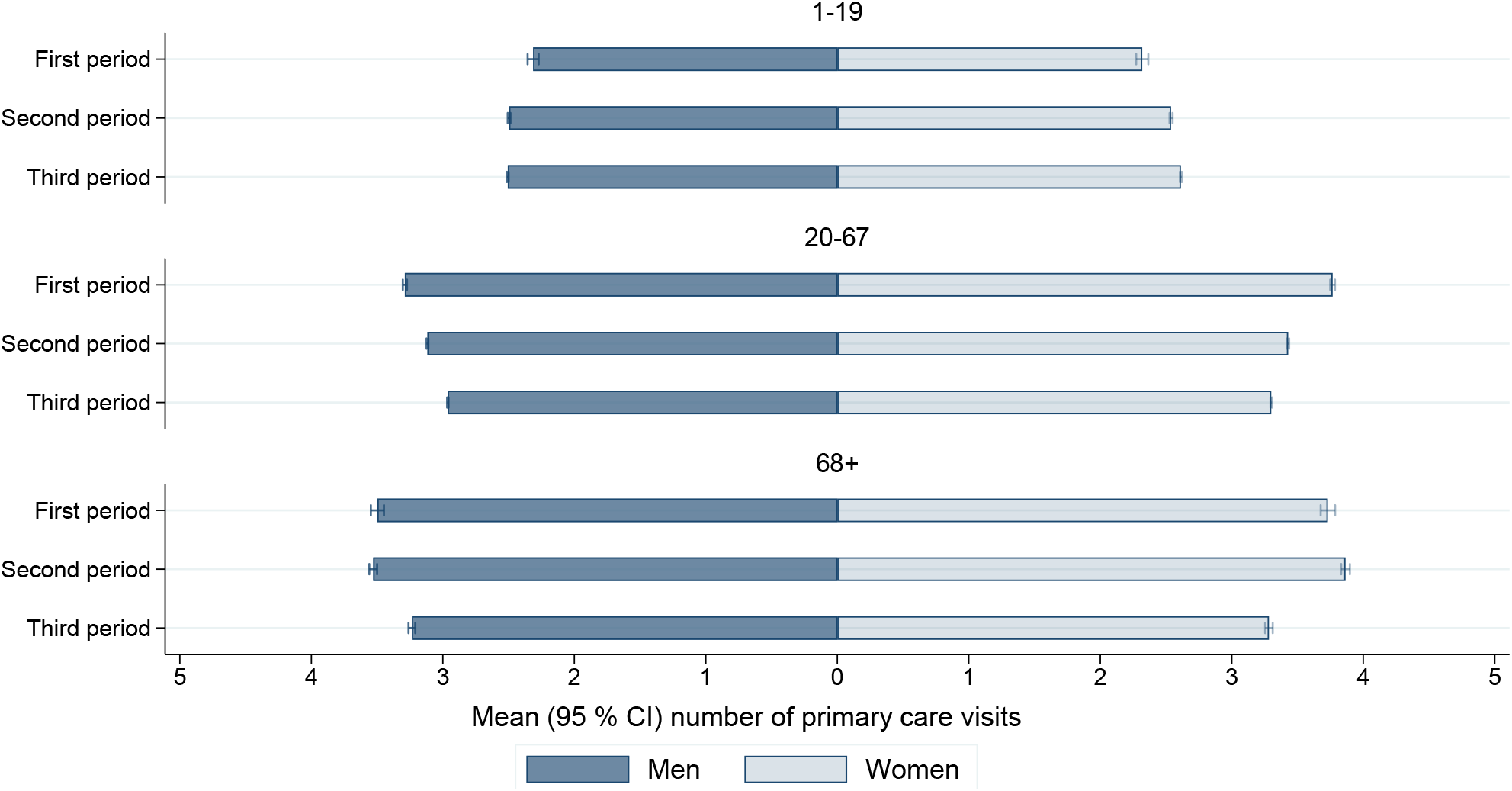

### Peak and total use of specialist care

We observed a significant shift in the total use of specialist care throughout the pandemic, from 14% being hospitalized at least once during the 1^st^ wave, compared to 4% during the 2^nd^ and 3^rd^ waves (Figure 4). During the 1^st^ wave, the share visiting specialist care peaked at the 2^nd^ and 3^rd^ day following a positive test (7%) (Figure 4). In contrast, and during the 2^nd^ and 3^rd^ wave, this share peaked at the 9^th^ and 10^th^ day following a positive test, with only 2% (Figure 4). S-Figure 4 shows that a minor proportion of children and adolescents, and a considerable proportion of elderly visited specialist care. Along this line, the mean number of bed-days was also longer for the middle-aged and elderly, than it was for children (Figure 5). Generally, women tended to have a lower number of bed-days than men (Figure 5). The mean number of bed-days tended to decrease from the 1^st^ to the 2^nd^ wave, before tending to increase again from the 2^nd^ to the 3^rd^ wave (Figure 5). However, the range of confidence intervals implied that the decrease was only statistically significant for men aged 20-67 and 68 or more (Figure 5). As expected, we see little changes in the cumulative share of persons visiting specialist care when excluding days –2 to 0, i.e. during the period in which any health care use could be assumed to be related to testing (S-Figure 5).

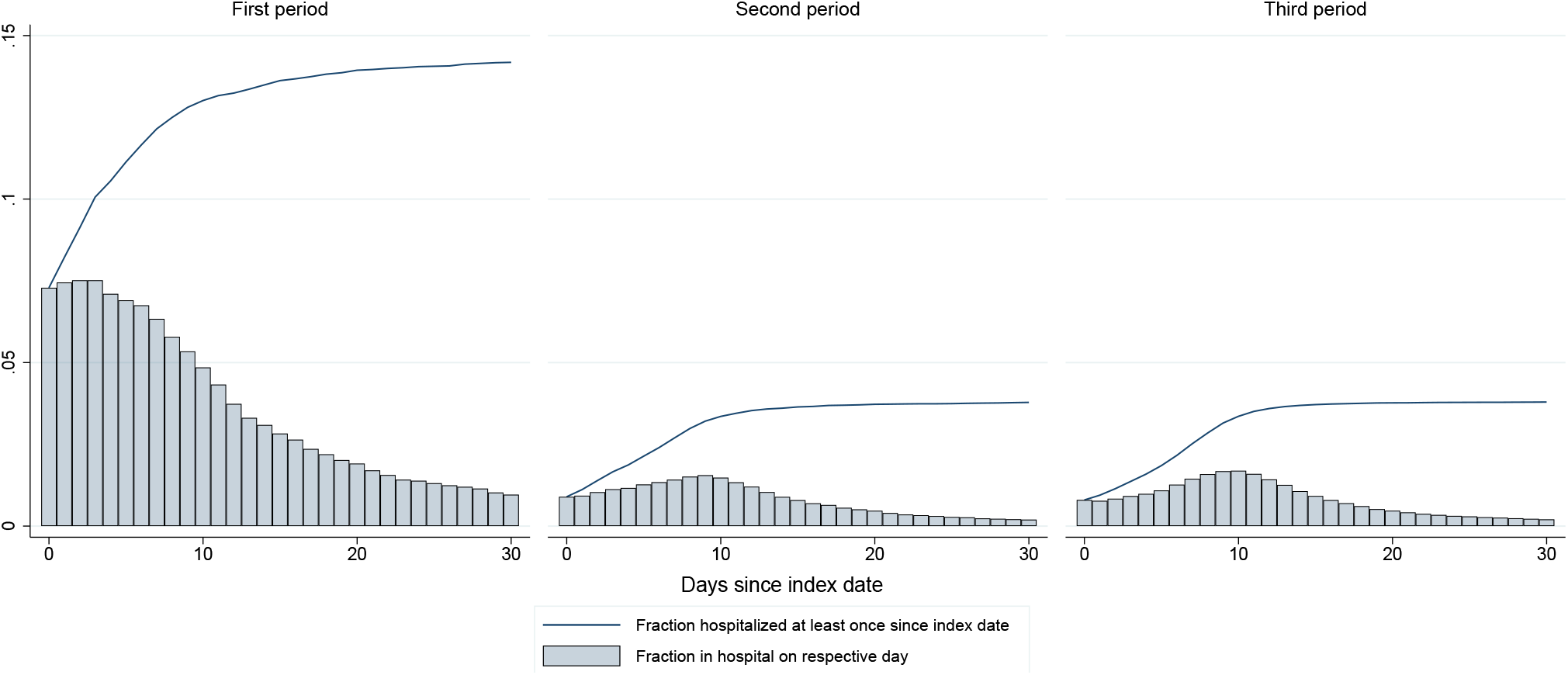

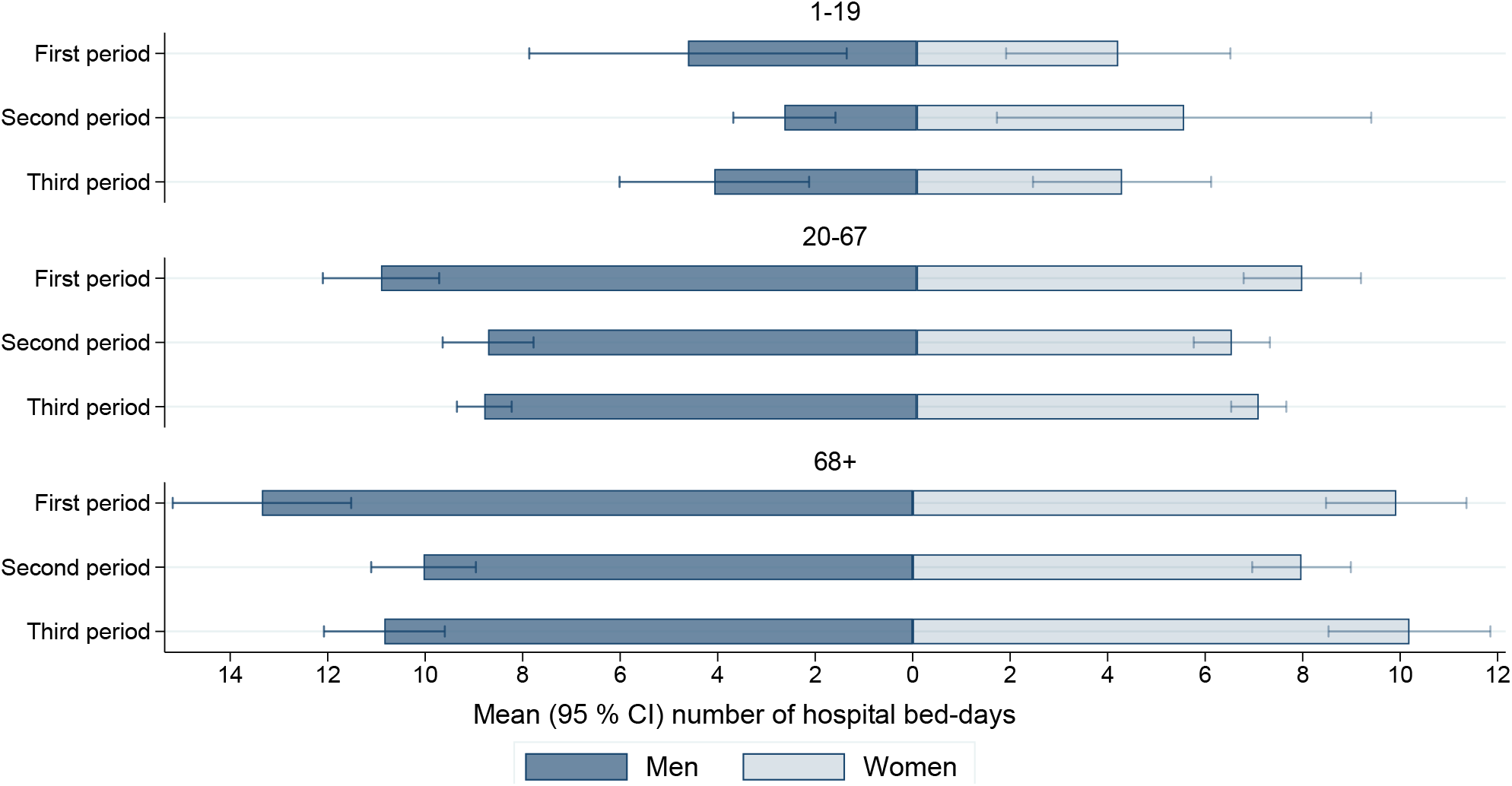

## Discussion

In this explorative study of all 122 699 persons testing positive for SARS-CoV-2 from March 1^st^ 2020 to May 31^st^ 2021 in Norway, we find that a high share of patients with COVID-19 (70-80%) has been treated exclusively in primary care in the acute phase, with a mean number of visits between 2 and 4. We also show that the acute 30-day impact of COVID-19 on specialist health care services decreased as the pandemic progressed: from 14% being hospitalized at least once during spring 2020, to 4% during spring 2021. This shift mainly applied to persons aged 20 or more.

An important strength of our study is that we could include everyone with a positive test throughout three major waves or periods of the pandemic. In this way, we could provide a comprehensive picture of *all* health care use following a positive PCR-test for these different waves, i.e. not restricted to specialist care as in previous studies [3, 4]. Moreover, we could provide details in primary and specialist care for different age and sex groups. We were unable to find a similar study for an effective comparison of our findings. However, it is evident from our data that the share using primary care is rather high for all age and sex groups, including children and adolescents (∼80%). In fact, whereas the mean number of primary care visits *decreased* for women and men aged 20 or more as the pandemic progressed, the same mean number *increased* for children and adolescents aged 1-19 years. Our findings imply that strengthening the clinical expertise and capacity of the primary care services (GPs and emergency wards) to handle COVID-19 patients may be important when facing future waves of the pandemic, and this may be particularly true for pediatric primary care. Causes for the increase in primary care use among children and adolescents are unknown, although it may partly be explained by differences in testing patterns as the pandemic progressed. Still, primary care use was utilized by almost the same share (∼70%) when excluding visits related to the testing and detection of COVID-19, and we suggest the increased primary care use among children and adolescents as a topic for future studies.

To our knowledge, the current study is the first to demonstrate the massive use of primary care services by COVID-19. This is important to report, given that a well-functioning primary care service is essential in reducing demands put on hospital services; it is essential to support rehabilitation of recovering patients; to improve palliative care; and sustain non-covid care [11]. As an example, S-Figure 1 shows that about 40% of persons aged 68 years or more who were hospitalized used primary care after discharge, and the share in this age group that could recover through help in primary care only increased from the 1^st^ to 2^nd^ wave (S-Figure 1). Still, in our study, the peak use of primary care was centered to the -2 to 0 days around positive test, implying that a certain proportion of the large amount of primary care visits took place in relation to testing and the detection of COVID-19. Indeed, when we excluded visits that were related to testing, the total share visiting primary care during the 30-day period decreased, yet only slightly (from ∼80% to ∼70%). Thus, the somewhat different patterns in primary care use from the 1^st^ to the 2^nd^ and 3^rd^ wave (Figure 2) may be explained by differences in testing criteria, i.e. the 1^st^ wave was the only wave with limited test availability and strict testing criteria (the elderly, persons at risk and health personnel).

Such differences in testing patterns are less likely to explain the decreasing demand put on specialist care services throughout the pandemic. Both the proportion hospitalized at least once, and the mean number of bed-days decreased from the 1^st^ to the 2^nd^ wave. Explanations for the decreased specialist care use may be related to a generally improved knowledge level of how COVID-19 behave clinically, both among the COVID-19 patients and among care personnel during the 2^nd^ and 3^rd^ waves of the pandemic. As an example, GPs may be hypothesized to more quickly refer to specialist care when the clinical knowledge was low and the risk of fatal outcomes high (as was the case when the pandemic stroke in March 2020). One example that may demonstrate this phenomenon is that the use of specialist care shifted from peaking on the 2^nd^-3^rd^ day during the 1^st^ wave, to peaking on the 9^th^-10^th^ days following positive test during the 2^nd^ and 3^rd^ wave (Figure 4). However, this observation might also reflect that testing occurred later in the course of the disease in the 1^st^ wave. Another important observation to our specialist care results, was the tendencies that the proportion hospitalized, as well as the mean number of bed-days in hospital *increased* from the 2^nd^ to the 3^rd^ wave (Figure 1, Figure 5). Although we did not aim to explore whether the severity of COVID-19 has changed, an important characteristic of the 3^rd^ wave of transmission has been the rise of mutant viruses. Recent reports are inconclusive as to whether mutant viruses result in more severe disease requiring more hospital care [10, 12, 13].

Several limitations should be mentioned. First, we do not know the causes or severity of complaints behind the care use following a positive test for SARS-CoV-2. Although we only included care visits with diagnostic codes of COVID-19, we could not separate the complaints affecting e.g. the respiratory or digestive system. Also, we had no comparison group, simply because we did not aim for any causal inference and because comparable data are not available for a similar epidemic or pandemic setting with other infectious diseases. However, in recent studies of post-acute COVID-19, we demonstrate a likely causal effect of being infected with SARS-CoV-2 on the post-acute health care use [14]. Here, we also exclusively included visits that were specific to COVID-19, i.e. we did not study all-cause visits. Second, our study was of an explorative and descriptive character. Thus, we looked for patterns and trends in a large amount of data using mainly graphs in a self-developed structure, such as the division of age into children and adolescents, adults in working age population and the elderly, and by sex. We did not apply any data-driven analyses in our exploration of pandemic trends in health care use, thus we might have missed important details. To combat some of these issues, we chose to present a large amount of raw data visualized as alluvial diagrams in the supplementary files (S-Figure 1). Third, we may have underestimated the care use among persons aged 68 years or more. Very frail persons live in care homes and receive institutionalized care that may not be registered in our data sources. And finally, and as mentioned above, we cannot exclude that some of our observations of changing (or stable) trends are due to differences in test criteria or -patterns as the pandemic progressed. Such patterns may differ across our groups of age and sex. However, if this is the case, the testing is obviously a part of health care use in relation to COVID-19, or else we would not have observed these visits. Thus, because testing for SARS-CoV-2 has been a part of the primary and specialist care services from the beginning of the pandemic, including care visits in relation to the detection of COVID-19 is still important in the public health question of whether the health services should be upscaled or downscaled in future similar situations.

In conclusion, we demonstrate a large acute impact of COVID-19 on primary care services, i.e. a stable proportion of around 70-80% visited primary care at least once during the first 30 days following a positive test. We also demonstrate a decreasing impact of COVID-19 on specialist care services from the 1^st^ to the 3^rd^ wave. These findings are important to report considering future waves of COVID-19, i.e. there may be a lower need for upscaling specialist care services and a large need for upscaling primary care services.

## Data Availability

Individual-level data of patients included in this manuscript after de-identification are considered sensitive and will not be shared. The study method and statistical analyses are all described in detail in the Methods and throughout the manuscript.

## Acknowledgements

We would like to thank the Norwegian Directorate of Health, in particular Director for Health Registries Olav Isak Sjøflot and his department, for excellent cooperation in establishing the emergency preparedness register. We would also like to thank Gutorm Høgåsen and Anja Elsrud Schou Lindman for their invaluable efforts in the work on the register. We would also like to thank Kjetil Telle, Anja Elsrud Schou Lindman, Karin Maria Nygård and Gunnar Øyvind Isaksson Rø for critically evaluating the content of the study. The interpretation and reporting of the data are the sole responsibility of the authors, and no endorsement by the register is intended or should be inferred. We would also like to thank everyone at the Norwegian Institute of Public Health who has been part of the outbreak investigation and response team.

## Code availability

Custom code in Stata is available upon request.

## Author contribution

Karin Magnusson designed the study. Fredrik Methi, Kjersti Helene Hernæs and Katrine Damgaard Skyrud had access to all the data in the study and takes full responsibility for the integrity of the data and the accuracy of the data analysis. Fredrik Methi, Kjersti Helene Hernæs and Katrine Damgaard performed the statistical analyses. Karin Magnusson drafted the study and critically evaluated all stages of the research process. All authors contributed with acquisition of data, conceptual design, analyses and interpretation of results. All authors contributed in drafting the article or critically revising it for important intellectual content. All authors gave final approval for the version to be submitted.

## References

1 WHO. WHO Director-General’s opening remarks at the media briefing on COVID-19 - 11 March 2020. https://www.who.int/dg/speeches/detail/who-director-general-s-opening-remarks-at-the-mediabriefing-on-covid-1911-march-2020. (Accessed 6 July 2021).

2 Guan WJ, Ni ZY, Hu Y, et al. Clinical characteristics of coronavirus disease 2019 in China. N Engl J Med 2020;382: 1708–20.

3 Richardson S, Hirsch JS, Narasimhan M, Crawford, JM, McGinn T, Davidson KW. Presenting characteristics, comorbidities, and outcomes among 5700 patients hospitalized with COVID-19 in the New York city area. JAMA 2020;323:2052–59.

4 Docherty AB, Harrison EM, Green CA, et al. Features of 20133 UK patients in hospital with covid-19 using the ISARIC WHO Clinical Characterisation Protocol: prospective observational cohort study. BMJ 2020;369:m1985.

5 Gutiérrez-Gutiérrez B, Del Toro MD, Borobia AM, et al. Identification and validation of clinical phenotypes with prognostic implications in patients admitted to hospital with COVID-19: a multicentre cohort study. The Lancet infectious diseases 2020;21:783–792.

6 Alexander GC, Tajanlangit M, Heyward J, Mansour O, Qato DM, Stafford RS. Use and content of primary care office-based vs telemedicine care visits during the COVID-19 pandemic in the US. JAMA network open 2020;3:e2021476–e2021476.

7 Norwegian Institute of Public Health. The Norwegian Emergency Preparedness Register (BEREDT C19), 2020. https://www.fhi.no/sv/smittsomme-sykdommer/corona/norsk-beredskapsregister-for-covid-19/

8 Norwegian Institute of Public Health. [Updates in Coronavirus Guidance]. 11.03.2020. https://www.fhi.no/nettpub/coronavirus/testing-og-oppfolging-av-smittede/testkriterier/?term=&h=1 (Accessed 6 July 2021).

9 Norwegian Institute of Public Health. [Testing criteria for SARS-CoV-2]. 08.02.2020 https://www.fhi.no/nettpub/coronavirus/testing-og-oppfolging-av-smittede/testkriterier/?term=&h=1 (Accessed 6 July 2021).

10 Norwegian Institute of Public Health. [New variants of SARS-CoV-2: Knowledge, risk and response: First update]. 12.01.2021. https://www.fhi.no/contentassets/c9e459cd7cc24991810a0d28d7803bd0/vedlegg/nye-varianter-av-sars-cov-2-kunnskap-risiko-og-respons-forste-oppdatering-13.01.2021.pdf (Accessed 7 July 2021).

11 Park S, Elliott J, Berlin A, Hamer-Hunt J, Haines A. Strengthening the UK primary care response to covid-19. BMJ 2020: 370.

12 Nagy A, Pongor S, Győrffy B. Different mutations in SARS-CoV-2 associate with severe and mild outcome, International Journal of Antimicrobial Agents, 2021:57:106272.

13 Whittaker R, Kristofferson AB, Seppälä E, et al. Trajectories of hospitalisation for patients infected with SARS-CoV-2 variant B.1.1.7 Norway, December 2020 – April 2021. medRxiv 2021.06.28.21259380v1 [Preprint]. 2021. https://doi.org/10.1101/2021.06.28.21259380

14 Skyrud KD, Telle KE, Magnusson K. Impacts of COVID-19 on long-term health and health care use. medRxiv 2021.02.16.21251807; doi: https://doi.org/10.1101/2021.02.16.21251807

