## Supplementary material for "Pandemic trends in health care use: From the hospital bed to the general practitioner with COVID-19": S-Figure

By Methi et al., 2021.


**S-Figure 1:** Alluvial diagram showing patient-flows in ten day-intervals by age group and period. p. 2

**S-Figure 2:** Fractions visiting primary care within 30 days of testing positive by age and sex. p. 3

**S-Figure 3:** Day by day cumulative fractions visiting primary care (GP or emergency ward) from one day after testing positive (day 1) to 30th day after positive test. p. 3

**S-Figure 4:** Fractions visiting specialist care within 30 days of testing positive by age and sex. p. 4

**S-Figure 5:** Day by day cumulative fractions visiting specialist care from one day after testing positive (day 1) to 30th day after positive test. p. 4

**S-Table 1:** Proportion dying during the first 30 days after positive test for SARS-CoV-2. p. 5

**S-Figure 1:** Alluvial diagram showing patient-flows in ten day-intervals by age group and period.

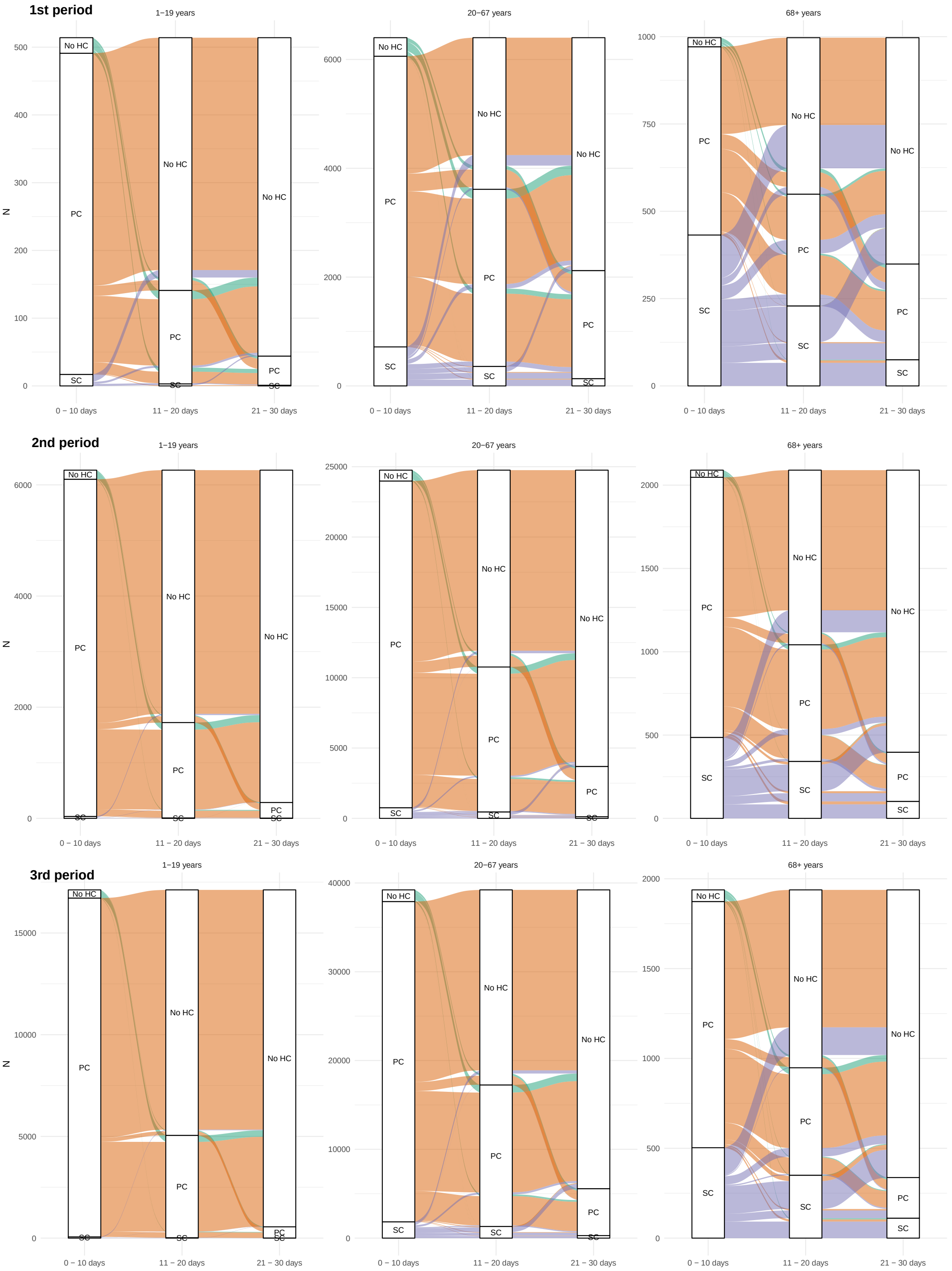


Note: No HC = No health care; PC = Primary care; SC = Specialist care. Green color shows persons with no health care use within the first 10 days of testing positive. Orange color shows persons with primary care within the first 10 days of testing positive. And purple color shows those in need of specialist care within the first 10 days of testing positive for SARS-CoV-2.

**S-Figure 2:** Fractions visiting primary care withing 30 days of testing positive by age and sex.

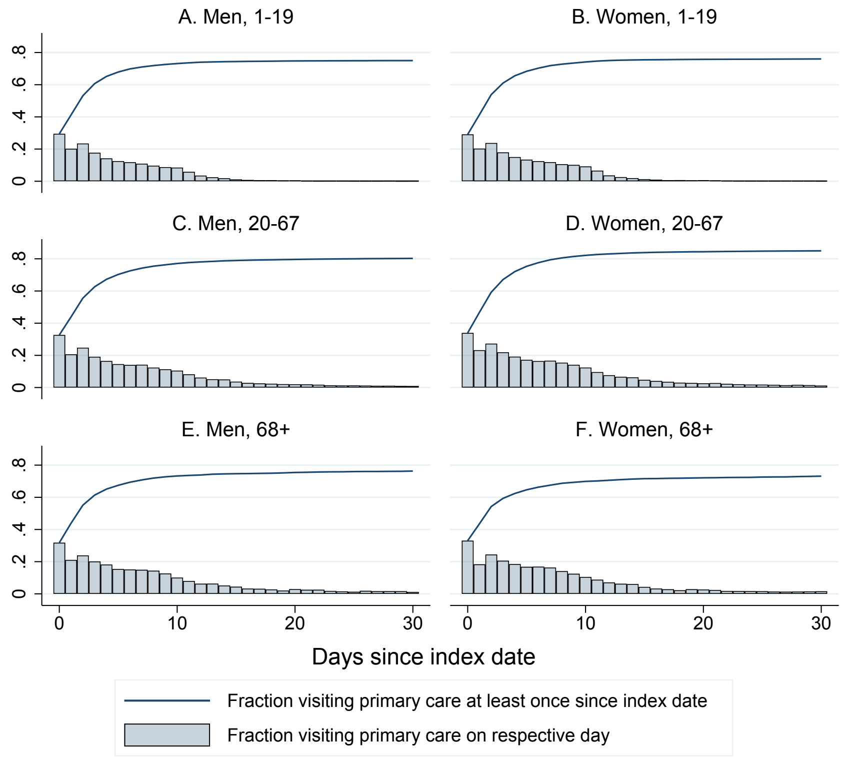


Note: Day 0 includes 0-2 days before testing positive.

**S-Figure 3:** Day by day cumulative fractions visiting primary care (GP or emergency ward) from one day after testing positive (day 1) to 30th day after positive test, i.e. excluding primary care visits that were related to the testing and detection of SARS-CoV-2.

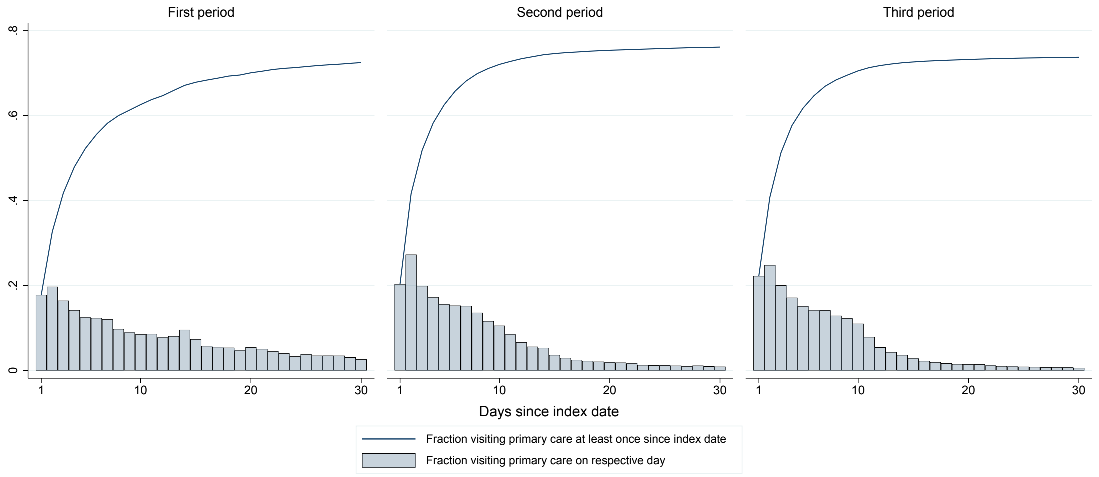

**S-Figure 4:** Fractions visiting specialist care withing 30 days of testing positive by age and sex.

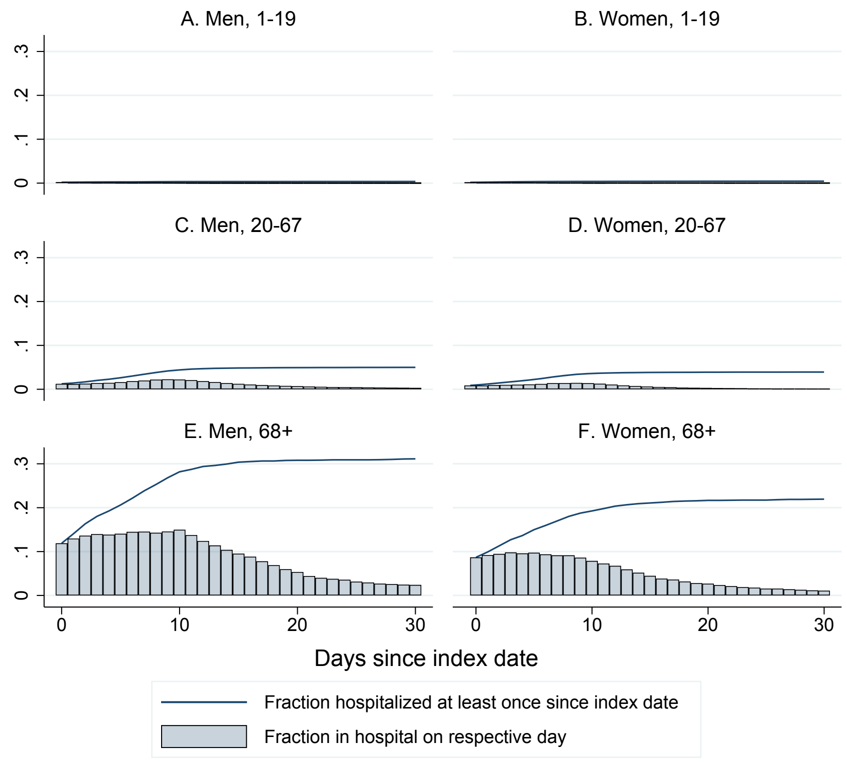


Note: Day 0 includes 0-2 days before testing positive.

**S-Figure 5:** Day by day cumulative fractions visiting specialist care from one day after testing positive (day 1) to 30th day after positive test, i.e. excluding specialist care visits that were related to the testing and detection of SARS-CoV-2.

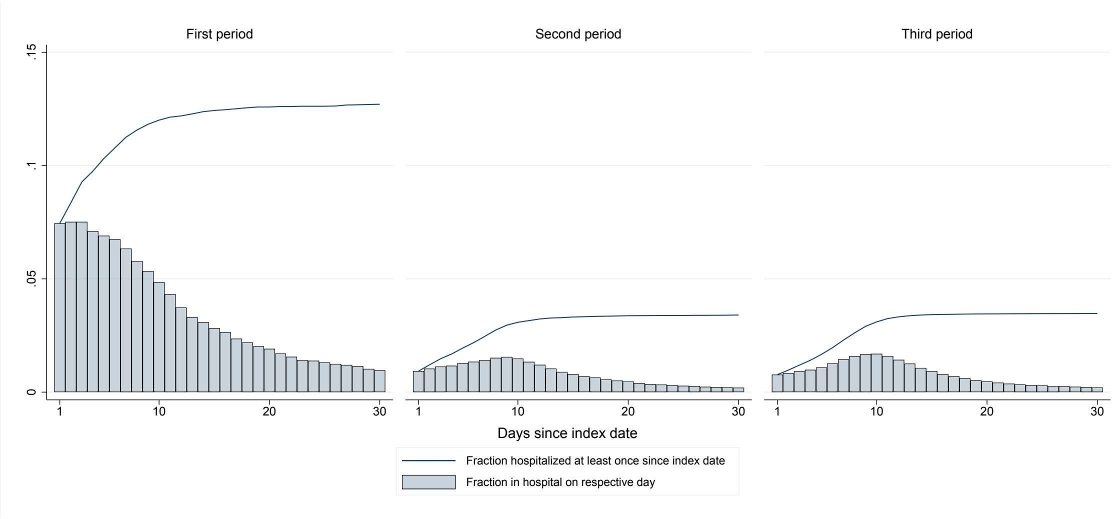

**S-Table 1:** Proportion dying during the first 30 days after positive test for SARS-CoV-2.

| | 1-19 years | | 20-67 years | | 68+ years | |
|---|---|---|---|---|---|---|
| | Men | Women | Men | Women | Men | Women |
| Whole sample | | | | | | |
| 1st period | 0 | 0 | 18 (0.5% [0.3%-0.8%]) | <5 | 110 (17.8% [15.0%-21.0%]) | 104 (15.9% [13.3%-18.9%]) |
| 2nd period | 0 | <5 | 22 (0.1% [0.1%-0.2%]) | 10 (0.1% [0.0%-0.1%]) | 99 (8.2% [6.8%-9.9%]) | 115 (8.6% [7.2%-10.2%]) |
| 3rd period | 0 | 0 | 33 (0.1% [0.1%-0.2%]) | 14 (0.1% [0.0%-0.1%]) | 99 (8.1% [6.7%-9.7%]) | 94 (7.8% [6.4%-9.4%]) |
| Pathway: No health care use | | | | | | |
| 1st period | 0 | 0 | <5 | 0 | 41 (35.7% [27.%-44.7%]) | 54 (33.1% [26.4%-40.7%]) |
| 2nd period | 0 | 0 | 6 (0.2% [0.1%-0.5%]) | <5 | 31 (16.2% [11.7%-22.1%]) | 49 (18.5% [14.3%-23.6%]) |
| 3rd period | 0 | 0 | <5 | <5 | 27 (12.8% [8.9%-18.0%]) | 30 (10.5% [7.4%-14.5%]) |
| Pathway: Primary care | | | | | | |
| 1st period | 0 | 0 | <5 | 0 | 15 (5.8% [3.6%-9.4%]) | 19 (6.5% [4.2%-9.9%]) |
| 2nd period | 0 | 0 | 5 (0.0% [0.0%-0.1%]) | <5 | 15 (2.1% [1.3%-3.4%]) | 33 (3.9% [2.8%-5.5%]) |
| 3rd period | 0 | 0 | 9 (0.0% [0.0%-0.1%]) | <5 | 11 (1.7% [0.9%-2.9%]) | 27 (3.8% [2.6%-5.5%]) |
| Pathway: Specialist care | | | | | | |
| 1st period | 0 | 0 | 15 (3.3% [2.0%-5.4%]) | <5 | 54 (22.0% [17.2%-27.5%]) | 31 (15.5% [11.1%-21.2%]) |
| 2nd period | 0 | <5 | 11 (2.3% [1.2%-4.0%]) | 8 (2.4% [1.2%-4.6%]) | 53 (17.8% [13.9%-22.6%]) | 33 (13.7% [9.9%-18.6%]) |
| 3rd period | 0 | 0 | 20 (1.6% [1.1%-2.5%]) | 11 (1.3% [0.8%-2.4%]) | 61 (17.5% [13.9%-21.8%]) | 37 (17.3% [12.8%-22.9%]) |

Note: 95% CI [in brackets] were calculated using the Wilson method.
